# Clinical benefits and adverse effects of genetically-elevated free testosterone levels: a Mendelian randomization analysis

**DOI:** 10.1101/19005132

**Authors:** Pedrum Mohammadi-Shemirani, Michael Chong, Marie Pigeyre, Robert W. Morton, Hertzel C. Gerstein, Guillaume Paré

## Abstract

**BACKGROUND:** Testosterone products are increasingly being prescribed to males for a variety of possible health benefits but the causal relationship between testosterone and health-related outcomes is unclear. Evidence from well-powered randomized controlled trials are difficult to obtain, particularly regarding effects on long-term or adverse outcomes. We sought to determine the effects of genetically-predicted calculated free testosterone (CFT) on 23 health outcomes.

**METHODS:** Genetic variants associated with CFT were determined from 136,531 white British males in the UK Biobank. One-sample and two-sample Mendelian randomization (MR) analyses were performed to infer the effects of genetically-predicted CFT on 23 health outcomes selected based on relevance with known or suspected effects of testosterone therapy.

**FINDINGS:** In males from the UK Biobank, 81 independent genetic variants were associated with CFT levels at genome-wide significance (p<5×10^−8^). Each 0.1 nmol/L increase in genetically-predicted CFT was associated with clinical benefits on increased heel bone mineral density (0.053 SD; 95% CI = 0.038 to 0.068; p=8.77×10^−12^) and decreased body fat percentage (−1.86%; 95% CI = −2.35 to −1.37; p=1.56×10^−13^), and adverse effects on increased risk of prostate cancer (OR=1.28; 95% CI=1.11 to 1.49; p=1.0×10^−3^), risk of androgenic alopecia (OR=1.82; 95% CI = 1.55 to 2.14; p=3.52×10^−13^), risk of benign prostate hyperplasia (BPH) (OR=1.81; 95% CI = 1.34 to 2.44; p=1.05×10^−4^) and hematocrit percentage (1.49%; 95% CI = 1.24 to 1.74; p=3.49×10^−32^).

**CONCLUSIONS:** Long-term elevated free testosterone levels cause prostate cancer, BPH, and hair loss while reducing body fat percentage and increasing bone density. It also has a neutral effect on type 2 diabetes, cardiovascular and cognitive outcomes. Well powered randomized trials are needed to address the effects of shorter term use of exogenous testosterone on these outcomes.

## INTRODUCTION

In developed countries, rising rates of both serum testosterone level testing and therapy initiation have been observed among older male patients^1,2^. In the USA, it is estimated 1.5-1.7% of males are currently prescribed testosterone^3,4^. Randomized clinical trials (RCT) have attempted to elucidate the benefits and risks of testosterone therapy^5,6^. These studies identified short-term beneficial effects on BMD^7^, sexual function^8^, body fat and muscle mass^9^, and anemia^10,11^; potential adverse effects on erythrocytosis^10,11^, venous thrombosis^12^ and coronary artery plaque^13^; and no effects on cognitive function^14^, liver fat^15^, fatigue^8^, or haemoglobin A1_c_ (HbA1_c_)^16^. However, given the logistic and financial challenges involved in a well-powered RCT with appropriate follow-up, there is unlikely to be satisfactory evidence regarding long-term effects and risks of adverse outcomes, such as myocardial infarction (MI), stroke and cancer^5^. Given the rates of testosterone prescription, efforts to resolve the causal effects of testosterone on health outcomes have important public health implications^6^.

Mendelian randomization (MR) is a technique for causal inference that leverages the random allocation of genetic variants to infer the unconfounded relationship between an exposure and outcome. Similar to the random assignment of participants to experimental groups in a RCT, genetic variants are randomly allocated at meiosis^17^. For instance, if individuals genetically randomized to produce higher testosterone develop different rates of cardiovascular disease (CVD), then MR analysis supports a causal effect of testosterone on risk of CVD (Supplementary Figure 1). Notably, this technique has previously replicated RCT findings, among others demonstrating causal roles for LDL cholesterol and dysglycemia on CVD risk^18,19^. Prior MR studies investigating the effects of testosterone have demonstrated harmful effects on lipid levels but inconsistent effects on CVD^20,21^. However, these studies have been limited by the small number of genetic variants and the use of total testosterone as opposed to free testosterone levels. Free testosterone is better representative of the effects of testosterone in the body, since total testosterone encompasses biologically inactive testosterone bound to transporter proteins^22^. Accordingly, studies have shown the main transporter of testosterone, sex hormone-binding globulin (SHBG), is a strong predictor of CVD risk factors independent of total testosterone and may be a source of confounding^23^. To date, there have been no MR studies investigating the effects of free testosterone.

We hypothesized that MR analyses would enable us to estimate the causal effects of longstanding exposure to high levels of free testosterone on selected health-related outcomes that have been previously investigated in randomized clinical trials of testosterone therapy. We first conducted a genome-wide association study (GWAS) for calculated free testosterone (CFT) in male participants of the UK Biobank (n = 136,531) cohort to identify genetic determinants of free testosterone levels. Then, using MR, we investigated the causal effects of lifelong genetically-elevated free testosterone levels on various health outcomes, encompassing: expected clinical benefits (physical activity, strength, fat-free body mass, body fat, BMD, dementia, depression) and potential adverse effects (androgenic alopecia, haematocrit, type 2 diabetes, liver fat, prostate cancer, blood pressure, CVD, heart failure, ischemic stroke) (Figure 1)^5–10,14–16,24^.

**Figure 1.**
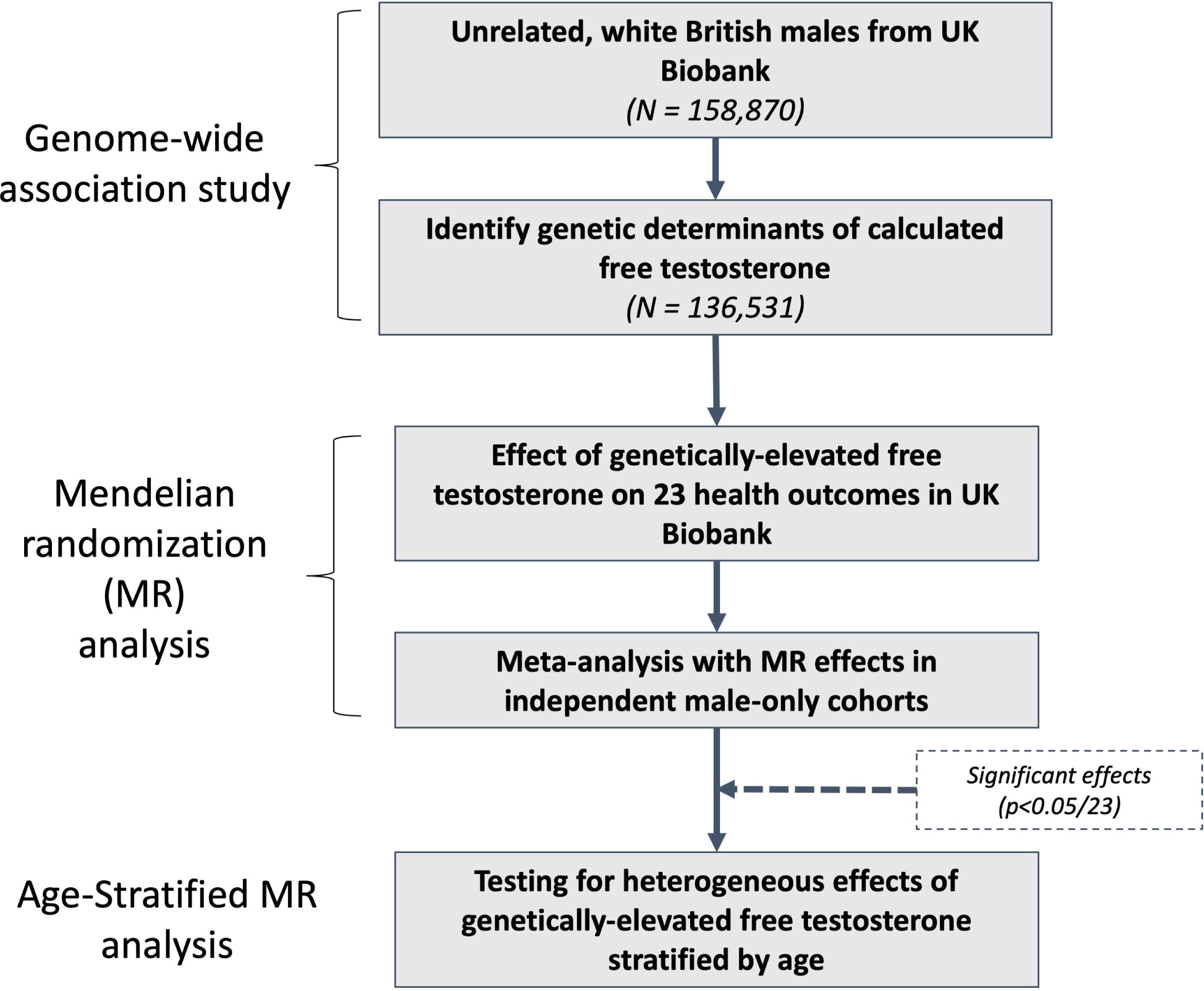
Flowchart depicting overall study design. Free testosterone levels were calculated in males from the UK Biobank cohort. Then, genetic variants were tested for association with levels of CFT and carried forward if: genome-wide significant (p<5×10^−8^) and unassociated with SHBG (p<0.05). These genetic variants represent genetically-predicted CFT (using variants unrelated to SHBG) and were tested for association with each of 23 health outcomes using Mendelian randomization. One-sample MR was performed using associations with 23 outcomes in males from the UK Biobank. Then, two-sample MR was performed using 10 dichotomous outcomes from independent cohorts with larger sample sizes. When male-specific summary statistics were available from independent cohorts, MR effect estimates were meta-analyzed. For statistically significant outcomes adjusting for multiple hypothesis testing (p<0.05/23), one-sample MR was repeated within age subgroups to identify differential effects in participants above versus below 65 years of age. CFT, calculated free testosterone; MR, Mendelian randomization; TT, total testosterone; SHBG, sex hormone-binding globulin

## METHODS

### Study Population - UK Biobank

The UK Biobank is a large-scale longitudinal cohort study that recruited over 500,000 people between the ages of 37-73 across the United Kingdom from 2006-2010^25^. Extensive health information was collected through self-reported medical histories and physical measures at recruitment, and ongoing health developments through linked electronic health records. UK Biobank received ethical approval from the North West Multi-Centre Research Ethics Committee (REC reference: 11/NW/0382). This research was conducted using the UK Biobank under Application Number 15255.

### Calculation of Free Testosterone

For our analyses, males in the UK Biobank were excluded if they had reported taking any androgen medication at recruitment based on field ID 20003 (n = 2,137). Free testosterone was calculated using the Vermeulen equation^22^ with levels of serum total testosterone, SHBG, and albumin measured at recruitment. CFT levels were winsorized such that outlying values greater or less than 4 standard deviations (SD) away from the mean in males were set to 4 SD.

### Genome-wide Association Study of Calculated Free Testosterone

For genome-wide association testing, UK Biobank samples were restricted to a subset of white British ancestry that were unrelated (greater than 3^rd^ degree). Genetic variants were restricted to those present in the Haplotype Reference Consortium panel^26^ with imputation quality greater than 0.6, no deviation from Hardy-Weinberg equilibrium (p > 1×10^−10^) and minor allele frequency greater than 0.1% (equivalent to approximately 150 copies of the minor allele in this male subset). After these exclusions, almost 12 million directly genotyped or imputed genetic variants were left in the model. BGENIE v1.2 ^27^ was used to run an additive genetic model in male participants from this subset of the UK Biobank. The model was adjusted for age, age^2^, chip type, assessment centre, and the first 20 genetic principal components.

Genetic variants near the *SHBG* gene have been shown to alter binding affinity for testosterone thereby violating assumptions of the Vermeulen equation^28^, and SHBG risks having pleiotropic effects through its binding of other sex hormones. Therefore, we tested genetic variants associated with CFT reaching genome-wide significance (GWS) (p ≤ 5×10^−8^) for evidence of association with natural log-transformed SHBG levels in the same subset of the UK Biobank (Supplementary Figure 2). Any genetic variants associated with SHBG (p<0.05) were removed. To arrive at an independent set of genetic variants, variants associated with CFT but not associated with SHBG were pruned based on linkage disequilibrium (LD) at a threshold of r^2^<0.01. The reference panel for LD was derived from 1000 Genomes Europeans phase 3 ^29^.

### Selection of Health-Related UK Biobank Outcomes

Thirty-one health outcomes were selected based on relevance with known or suspected effects of testosterone therapy, and they were classified based on expected beneficial or adverse effects from randomized clinical trial data. Outcomes with expected beneficial effects were fractures at any site, heel BMD, body fat percentage, dementia, depression, erectile dysfunction, handgrip strength, physical activity level measured by wrist-worn accelerometer, and whole body fat-free mass. Outcomes with potential adverse effects were stroke, androgenic alopecia, arterial embolism and thrombosis, benign prostate hyperplasia (BPH), blood pressure, glucose, haematocrit percentage, haemoglobin A1c, heart failure, liver fat, prostate cancer, MI, testicular cancer, type 2 diabetes (T2D), and venous thromboembolism. Detailed descriptions of all outcomes are shown in Supplementary Table 1 and Supplementary Methods. Given the small number of cases for several outcomes, power calculations were performed for MR analyses using a significance level of 0.05, and any outcomes with detectable odds ratios less than 0.5 or greater than 2 per 0.1 nmol/L at 80% power were excluded (Supplementary Table 2)^30^. After these exclusions, there were 23 health outcomes that remained for subsequent MR analyses in this study.

The association of all independent genetic variants associated with CFT were determined for each of the 23 outcomes using additive genetic models in BGENIE v1.2 ^27^ and adjusted for the same covariates as the model for CFT. For dichotomous outcomes, odds ratios were approximated as previously described^31^ by converting linear effect estimates to log-odds scale using: 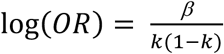, where *k* is the proportion of cases for the given outcome

### Mendelian randomization – One-sample design

For each of the 23 outcomes, one-sample MR analysis was used to combine the effect of each independent genetic variant on CFT with its effect on the outcome. Weighted median method was used to infer the causal effect of CFT on the outcome, which selects the median causal estimate after weighing estimates of each individual genetic variant according to their variance^32^. The resulting estimate is robust to horizontal pleiotropy when at least 50% of weights belong to valid genetic variants^32^. Given the polygenic nature of testosterone and potential for pleiotropy, this method was selected to ensure reliable estimates. Outcomes were deemed statistically significant after adjusting for multiple hypothesis testing of 23 health outcomes using Bonferroni correction (p<2.17×10^−3^). Effect estimates were reported per 0.1 nmol/L increase in CFT levels based on approximate changes in response to testosterone therapy^33,34^.

Sensitivity analyses ensured effects weren’t driven by any single variant. For statistically significant outcomes, leave-one-out analyses was performed where MR analysis was repeated after each genetic variant was excluded. Furthermore, genetic variants were assessed for “weak instrument bias”, which can result in biased estimates if genetic variants don’t explain enough variance in CFT levels^35^. The F-statistic was 4952 for the genetic variants used in this MR, which was considered a strong instrument based on the recommended threshold of greater than 10 ^35^.

### Mendelian randomization – Two-sample design

Given the limited number of cases in the UK Biobank for dichotomous outcomes, the data were supplemented using results from 10 publicly-available studies published elsewhere. Data from these independent studies were analyzed using a two-sample MR approach with the 81 genetic variants associated with CFT from the UK Biobank, and the association of each CFT-associated genetic variant with the outcome was obtained from the published GWAS. Generally, two-sample MR has the advantage of enabling the use of larger sample sizes and biases estimates towards the null under weak instrument conditions. We identified 10 publicly available GWAS summary statistics corresponding to dichotomous outcomes used in the one-sample MR analysis (Supplementary Table 3)^36–41^. For each outcome, MR analysis was similarly performed using the weighted median method. For male-specific outcomes, MR effect estimates from two-sample and one-sample analyses were meta-analysed under a fixed effects model using the *meta* package in R^42^.

### Mendelian randomization - Age-stratified

Given age-related decline in testosterone levels and selection criteria in RCTs, we stratified the UK Biobank male participants into two groups based on age at recruitment above or below 65 years to investigate differential effects of free testosterone levels in these age groups. For significant outcomes from the MR analysis (p<2.17×10^−3^), one-sample MR analysis was repeated as described previously separately in each subgroup to interrogate the effect of CFT on the given outcome according to age. P-values for heterogeneity between age groups were calculated under a fixed effects model in the *meta* package.

All MR analyses were implemented using the *TwoSampleMR* package^43^. All statistical analyses were performed under R version 3.6.0, unless otherwise specified. A two-sided p-value less than 5×10^−8^ for GWAS, 2.17×10^−3^ (0.05/23 outcomes) for standard MR analyses, and 0.05 for heterogeneity in age-stratified MR analyses was considered statistically significant.

### Patient and public involvement

This research was done without patient involvement. Patients were not invited to comment on the study design and were not consulted to develop patient relevant outcomes or interpret the results. Patients were not invited to contribute to the writing or editing of this document for readability or accuracy.

## RESULTS

### Genetic determinants of calculated free testosterone in males

There were 158,870 males in the unrelated white, British subset of the UK Biobank cohort. To calculate free testosterone levels, we excluded individuals that had missing levels of total testosterone, SHBG and albumin, or self-reported taking androgen medications. After these exclusions, the study population consisted of 136,531 males with an average CFT of 0.21 nmol/L (Supplementary Table 4 and Supplementary Figure 3).

There were 9,150 genetic variants associated with CFT that reached GWS (p<5×10^−8^) (Supplementary Figure 4). After removing genetic variants associated with natural-log transformed SHBG and keeping only independent signals, there were 81 genetic variants carried forward for subsequent genetic analyses (Supplementary Table 5). Altogether, these 81 genetic variants explained 3.4% of the total variance of CFT levels in males from the UK Biobank.

### Effect of free testosterone on 23 health outcomes using Mendelian randomization analysis

In males from the UK Biobank, sample size for the quantitative risk factors ranged from 1,595 to 156,403, while number of cases for dichotomous outcomes ranged from 1,003 to 70,283 (Table 1). After adjusting for the 23 outcomes tested, one-sample MR analysis identified significant effects of CFT on haematocrit percentage, body fat percentage, heel BMD, androgenic alopecia, and BPH (Table 1). Each 0.1 nmol/L higher CFT had beneficial effects on increased heel BMD (0.053 SD; 95% CI = 0.038 to 0.068; p=8.77×10^−12^) and decreased body fat percentage (−1.86%; 95% CI = −2.35 to −1.37; p=1.56×10^−13^), but deleterious effects on increased haematocrit percentage (1.49%; 95% CI = 1.24 to 1.74; p=3.49×10^−32^), risk of androgenic alopecia (OR=1.82; 95% CI = 1.55 to 2.14; p=3.52×10^−13^) and risk of BPH (OR=1.81; 95% CI = 1.34 to 2.44; p=1.05×10^−4^). Leave-one-out analyses did not identify any outlying individual genetic variants responsible for the observed effects on any significant outcomes.

**Table 1.** Effect of calculated free testosterone on 23 health outcomes in males from the UK Biobank

| Outcome | Effect per 0.1 nmol/L increased CFT<br>(95%CI) | P-value | Sample Size<br>Cases/Controls |
| --- | --- | --- | --- |
| <u>Outcomes with Expected Clinical Benefits</u> |  |  |  |
| **Body Fat Percentage** | **-1.86 % (-2.35 to -1.37)** | **1.56E-13** | **154095** |
| **Heel Bone Mineral Density** | **0.053 SD (0.038 to 0.068)** | **8.77E-12** | **90597** |
| Whole Body Fat-Free Mass | 0.92 kg (0.27 to 1.58) | 5.67E-03 | 154262 |
| Handgrip Strength | 0.61 kg (-0.06 to 1.28) | 0.074 | 156403 |
| Accelerometer-based Physical Activity | 0.90 milligravity (-0.43 to 2.23) | 0.19 | 30439 |
| All Fracture | OR = 0.82 (0.6 to 1.12) | 0.21 | 9133/148098 |
| Depression | OR = 1.28 (0.84 to 1.95) | 0.26 | 4725/152506 |
| All Dementia | OR = 0.86 (0.36 to 2.1) | 0.75 | 1003/156228 |
| <u>Outcomes with Potential Adverse Effects</u> |  |  |  |
| **Hematocrit Percentage** | **1.49 % (1.24 to 1.74)** | **3.49E-32** | **152893** |
| **Androgenic Alopecia** | **OR = 1.82 (1.55 to 2.14)** | **3.52E-13** | **70283/85757** |
| **Benign Prostatic Hyperplasia** | **OR = 1.81 (1.34 to 2.44)** | **1.05E-04** | **10894/146337** |
| Prostate Cancer | OR = 1.58 (1.13 to 2.2) | 7.71E-03 | 7586/149645 |
| Myocardial Infarction | OR = 1.45 (1.08 to 1.95) | 0.013 | 9398/147833 |
| Hemoglobin A1c | -0.47 mmol/mol (-1.02 to 0.08) | 0.096 | 149829 |
| Diastolic Blood Pressure | 0.54 mmHg (-0.21 to 1.29) | 0.16 | 143809 |
| All Stroke | OR = 1.32 (0.87 to 2.02) | 0.19 | 4569/152662 |
| Venous Thromboembolism | OR = 0.78 (0.5 to 1.22) | 0.28 | 4127/153104 |
| Type 2 Diabetes | OR = 1.07 (0.8 to 1.43) | 0.65 | 11079/146152 |
| Glucose | -0.02 mmol/L (-0.13 to 0.09) | 0.68 | 138308 |
| Heart Failure | OR = 1.09 (0.7 to 1.69) | 0.71 | 4288/152943 |
| Ischemic Stroke | OR = 0.91 (0.51 to 1.62) | 0.75 | 2122/155109 |
| Liver Fat | -0.50 % (-3.80 to 2.80) | 0.77 | 1595 |
| Systolic Blood Pressure | -0.18 mmHg (-1.55 to 1.20) | 0.80 | 143805 |

**Bolded** rows are significant adjusting for multiple hypothesis testing of 23 outcomes using Bonferroni correction ( $p < 2.17 \times 10^{-3}$ ) CFT, calculated free testosterone

To increase the statistical power of the MR analyses, two-sample MR analysis was performed for dichotomous outcomes using independent summary statistics from GWAS with greater numbers of cases. Male-specific summary statistics were available for prostate cancer from the Prostate Cancer Association Group to Investigate Cancer-Associated Alterations in the Genome (PRACTICAL) consortium, and combined male and female summary statistics for 10 other dichotomous outcomes (Supplementary Table 3). The effect of CFT was directionally consistent with the one-sample MR analysis for prostate cancer. Given no overlap between the samples in the UK Biobank and PRACTICAL consortium summary statistics, effect estimates were meta-analysed and reached statistical significance (OR=1.28; 95% CI=1.11 to 1.49; p=1.0×10^−3^). Similarly, there was a consistent null effect observed for ischemic stroke and other stroke subtypes, Alzheimer’s disease, and T2D (Supplementary Table 6), but these effect estimates were not meta-analysed due to lack of available male-specific summary statistics.

### Differential effects of free testosterone on health outcomes according to age subgroups

To identify differential effects of CFT between age groups, we repeated MR analysis in males from the UK Biobank stratified according to participant’s age below or above 65 years. Age-specific effects of CFT were then determined for statistically significant (p<2.17×10^−3^) outcomes from MR analyses. Genetically-elevated CFT showed a risk-conferring effect on BPH (OR = 1.80; 95% CI= 1.22 to 2.65; p=2.89×10^−3^) and prostate cancer (OR = 2.02; 95% CI= 1.29 to 3.65; p=2.10×10^−3^) specific to males under 65 years of age (p_heterogeneity_= 0.019 for BPH and 0.038 for prostate cancer) (Supplementary Figure 5). Androgenic alopecia, haematocrit percentage, body fat percentage, and heel BMD showed no significantly different effect in participants above versus below 65 years of age.

## DISCUSSION

We herein perform an MR analysis of CFT to identify effects of elevated endogenous free testosterone in males on 23 health outcomes. MR analyses demonstrated that each 0.1 nmol/L increase in CFT was associated with adverse effects on increased risk of prostate cancer, risk of androgenic alopecia, risk of BPH, and hematocrit percentage, but beneficial effects on increased heel BMD and decreased body fat percentage. These findings are consistent with short-term effects in randomized trials of testosterone therapy, and opposite effects of 5α-reductase inhibitors – inhibiting the conversion of testosterone into the more potent dihydrotestosterone – as a treatment for androgenic alopecia and BPH^6,44,45^. Furthermore, MR analyses showed no effect of free testosterone on several hard endpoints, such as dementia, MI, stroke, fractures, and T2D. Accordingly, these results cast doubt on claims of cardiovascular risk or benefit for testosterone therapy^46^.

After meta-analysis of MR results from the UK Biobank and PRACTICAL consortium studies, there was a statistically significant risk-conferring effect of free testosterone on prostate cancer (p=1.0×10^−3^). Prostate cancer is the leading cause of cancer and second leading cause of cancer death among men in the USA^47^. Androgen suppression has long been used as treatment for both BPH^48^ and prostate cancer^45^. However, there is inconclusive RCT evidence regarding the effects of elevated testosterone via either endogenous or exogenous means on development of BPH or prostate cancer^49^. These MR results suggest 1.3-fold increased risk of prostate cancer per 0.1 nmol/L increase in free testosterone levels, which is comparable to changes observed after initiation of testosterone therapy^33,34^. Moreover, age-stratified analyses provided suggestive evidence of a specific effect on BPH and prostate cancer in individuals under 65 years of age. These findings warrant further investigation in clinical trials of exogenous testosterone supplementation and calls for greater scrutiny in this patient population.

There are several limitations of this study. First, an assumption of the MR analysis is that the effect of the genetic variant on the outcome occurs only through free testosterone levels, such that there are no pleiotropic effects through other proteins or mechanisms^17^. This concern was minimized by the use of multiple genetic variants, which limited the likelihood of a common alternative pathway confounding our observation. Moreover, we performed standard sensitivity analyses and excluded genetic variants associated with SHBG levels, which is a potential source of pleiotropy through its effects on other hormones. Additionally, one-sample MR may be susceptible to bias towards the confounded estimate if the genetic variants are “weak instruments”, which can occur if the genetic variants don’t explain enough of the variance in free testosterone levels^35^. To address this concern, we confirmed the selected genetic variants were strong instruments using a common threshold in MR literature (F-statistic > 10)^35^. Furthermore, two-sample MR analyses afforded larger sample sizes for certain dichotomous outcomes, and yielded a risk-conferring effect of CFT on prostate cancer and nonsignificant effects on Alzheimer’s disease, coronary heart disease, stroke subtypes, and T2D. However, besides prostate cancer, these results should be interpreted with caution as these GWAS summary statistics didn’t provide sex-specific genetic estimates (Supplementary Table 3). Consequently, if the effect of CFT genetic associations are male-specific, the causal effect of free testosterone in males may have been diluted or counteracted by the inclusion of females. Finally, these MR results represent lifelong effects of endogenous free testosterone and may not necessarily reflect effects of exogenous testosterone therapy, which can vary in duration, age of initiation, and dosage.

Taken altogether, the decision to initiate long-term testosterone use warrants careful consideration of benefits and risk. From a lifestyle perspective, beneficial effects on body fat percentage and sexual function should be weighed against detrimental effects on androgenic alopecia and adverse urinary symptoms arising from BPH. From a clinical perspective, there may be a beneficial effect on hematocrit percentage and BMD but no significant effects on hard clinical endpoints (e.g., CVD, fracture, and T2D) except a risk-conferring effect on prostate cancer with evidence of a possible effect specific to younger males. Ultimately, well-designed and appropriately powered RCTs, such as the ongoing TRAVERSE trials (clinicaltrials.gov, NCT03518034), are necessary to conclusively address questions of safety and effectiveness of testosterone therapy. However, as demonstrated in this study, genetically-informed analyses can be powerful tools to aid health professionals in prioritizing allocation of limited resources towards investigating the most pressing questions.

## Data Availability

Associations of genetic variants with calculated free testosterone in the UK Biobank are provided in the supplementary data. All other UK Biobank data is available by application directly to the UK Biobank. All other genome-wide association study summary statistics are publicly-available from the corresponding consortia: CARDIoGRAMplusC4D (http://www.cardiogramplusc4d.org/data-downloads/), DIAGRAM (https://www.diagram-consortium.org/downloads.html), IGAP (http://web.pasteur-lille.fr/en/recherche/u744/igap/igap_download.php), MEGASTROKE (http://www.megastroke.org/download.html), and PRACTICAL (http://practical.icr.ac.uk/blog/?page_id=8164).

https://www.diagram-consortium.org/downloads.html

http://web.pasteur-lille.fr/en/recherche/u744/igap/igap_download.php

http://www.megastroke.org/download.html

http://practical.icr.ac.uk/blog/?page_id=8164

## ACKNOWLEDGEMENTS

The authors are thankful for all the participants that contributed to the UK Biobank study. Also, the authors would like to thank the CARDIoGRAMplusC4D, DIAGRAM, IGAP, MEGASTROKE, and PRACTICAL consortia for making summary statistics publicly-available for this work.

